# How the COVID-19 pandemic and cost-of-living crisis shaped reach and engagement in the ECAIL trial targeting socially disadvantaged families: an interdisciplinary implementation study

**DOI:** 10.64898/2026.05.14.26353230

**Authors:** Delphine Poquet, Camille Le Gal, Pascale Hincker, Laurent Béghin, Dominique Deplanque, Damien Subtil, Oriane Sion, Benjamin Cavalli, Lucie Vanhoutte, Victoria Jacobsen, Ketevan Maar, Ioannis Sakellaris, Blandine de Lauzon Guillain, Marie-Aline Charles, Delphine Ley, Priscille Sauvegrain, Sandrine Lioret

## Abstract

**Background:** The ECAIL trial, launched in 2017, targets hard-to-reach families and evaluates a multicomponent childhood obesity prevention intervention. At a maternity hospital in Lille, France, healthcare providers screened pregnant women experiencing social vulnerability, and dietitians delivered a home-based intervention until age 2. The COVID-19 pandemic led to a six-month suspension in 2020. This study compared eligibility and participation before the pandemic and after resumption, and examined how the pandemic and subsequent cost-of-living crisis shaped implementation and reach.

**Methods:** We analyzed 5,744 eligibility questionnaires distributed at the maternity ward. Inclusion criteria included ≥1 indicator of social vulnerability (e.g., socioeconomic disadvantage, precarious housing, or social isolation). To capture implementation experiences, a psychosocial researcher conducted a focus group with six dietitians delivering the intervention; it was recorded, transcribed, and analyzed thematically focusing on reach, acceptability, and adaptation.

**Results:** Eligibility increased from 29.7% (n=955) prepandemic to 33.6% (n=849) after resumption, while the distribution of vulnerability criteria remained similar across periods: 78.3% received social/medical benefits; employment was not the main source of household income for 58.7%; 24.4% experienced financial hardship; 14.7% reported social isolation; 6.0% lived in precarious housing; and 19.0% had three or more vulnerabilities. Participation among eligible women remained stable (24.6%; n=443). Qualitative findings indicated dietitians’ satisfaction and participants’ enthusiasm for the resumption of home visits, particularly in addressing social isolation. After resumption, the introduction of a pre-visit COVID-19 questionnaire reduced missed appointments. Converging qualitative and quantitative findings indicated sustained, and in some cases strengthened, provider engagement despite pandemic-related strain on hospital services.

**Conclusions:** This study shows that a complex intervention can maintain reach and acceptability through adaptive implementation under major contextual disruptions. The rapid resumption of home-based services emerged as a robust strategy for engaging and retaining socially disadvantaged families, highlighting the importance of flexible, context-sensitive approaches during social and economic crises.

**Trial registration:** Clinicaltrials.gov NCT03003117; registration date: 21/12/2016; https://clinicaltrials.gov/study/NCT03003117

## Introduction

In 2020, the COVID-19 pandemic triggered a massive health crisis worldwide, accompanied by prolonged economic and social repercussions. Following the initial lifting of pandemic-related health restrictions and in the context of subsequent geopolitical shocks, including the invasion of Ukraine, energy and food prices surged across the Eurozone (1). Although governments implemented exceptional measures, this inflation reduced consumer purchasing power, especially among low-income households (2). In France, for example, according to the National Institute of Statistics and Economic Studies (Insee), the cost of the “food basket” rose more sharply for lower income households. This was partly because items like oils and other fatty products, representing a larger share of their food spending, were among the products most affected by inflation, with prices rising by about 30% in January 2023 compared with the previous year (3). Likewise, because transportation and housing make up a larger share of their consumption basket, these households were also more severely affected by rising energy prices (4). As a result, low-income households, who were already vulnerable before the shocks, were and continue to be disproportionately affected by these successive health and cost-of-living crises (5), illustrating a cumulative disadvantage often described as the poverty penalty (6). Moreover, the rise in poverty and food insecurity following the COVID-19 pandemic (7, 8) has further exacerbated social, economic, and health inequalities (9, 10).

Numerous studies have underscored the detrimental impact of the COVID-19 pandemic on health-related behaviors (such as physical activity, sedentary behaviors, and dietary intake), as well as on obesity and mental health in both adults and children (11–17). Far less attention, however, has been given to how such large-scale disruptions have affected the implementation of ongoing public health interventions, particularly those targeting socially disadvantaged populations. From an implementation science perspective, understanding how implementation outcomes such as reach, engagement, and fidelity are sustained, adapted, or disrupted under crisis conditions remains a critical gap in the literature (18). Frameworks such as RE-AIM highlight the importance of examining reach, adoption, and implementation processes alongside effectiveness, particularly for interventions targeting socially disadvantaged groups (19, 20) that are considered “hard-to-survey”, that is, populations who are not only difficult to sample, but also challenging to identify, reach, recruit, engage, and retain over time (21, 22).

We seek to address this gap by analyzing quantitative and qualitative data collected within the ongoing prEgnanCy and eArly Childhood nutrItion triaL (ECAIL), launched in 2017 in northern France (23). The overarching aim of ECAIL is to assess the effectiveness of a multilevel and multibehavioral intervention initiated during pregnancy that combines nutritional counseling with facilitated access to healthy foods at a reduced price, with the goal of promoting healthy lifestyle behaviors and preventing obesity in young children from underserved families. Building on an interdisciplinary and implementation-focused approach, the present study aims to (i) describe and compare eligibility and participation rates before the pandemic and after the trial resumed six months later, with particular attention to reach and its social patterning, and (ii) explore how the pandemic and subsequent cost-of-living crisis shaped the implementation of ECAIL, including adaptations to delivery and stakeholder engagement in the trial.

## Methods

### Study design and participants

ECAIL is an ongoing single-blind randomized controlled trial led by the French national institute for health and medical research (Inserm) and underway since 2017 in northern France to evaluate the effectiveness of an existing program, named *Malin* (24), delivered by trained dietitians. Beyond its evaluative design, ECAIL is also a participatory action research project (25), co-created with members of the *Malin* nonprofit association and its historical stakeholders, namely the French Red Cross and two French pediatric societies (the French Society of Pediatrics and the French Association for Ambulatory Pediatrics), in close collaboration with investigators from the Lille University Hospital and the Valenciennes Hospital. This co-creation process aimed to enhance the intervention’s contextual relevance, feasibility, and acceptability for socially disadvantaged families, as well as its integration into existing care pathways. More details about the ECAIL trial design, intervention components, and implementation procedures are provided in the published protocol (23).

Pregnant women were screened for eligibility during routine prenatal care at the maternity wards of Lille University Hospital (from March 2017 to April 2023; **Fig 1**), where 443 women were enrolled, and at Valenciennes Hospital. The present analysis focuses exclusively on the Lille site, as the Valenciennes center opened in June 2021, more than one year after the onset of COVID-19.

**Fig 1.**
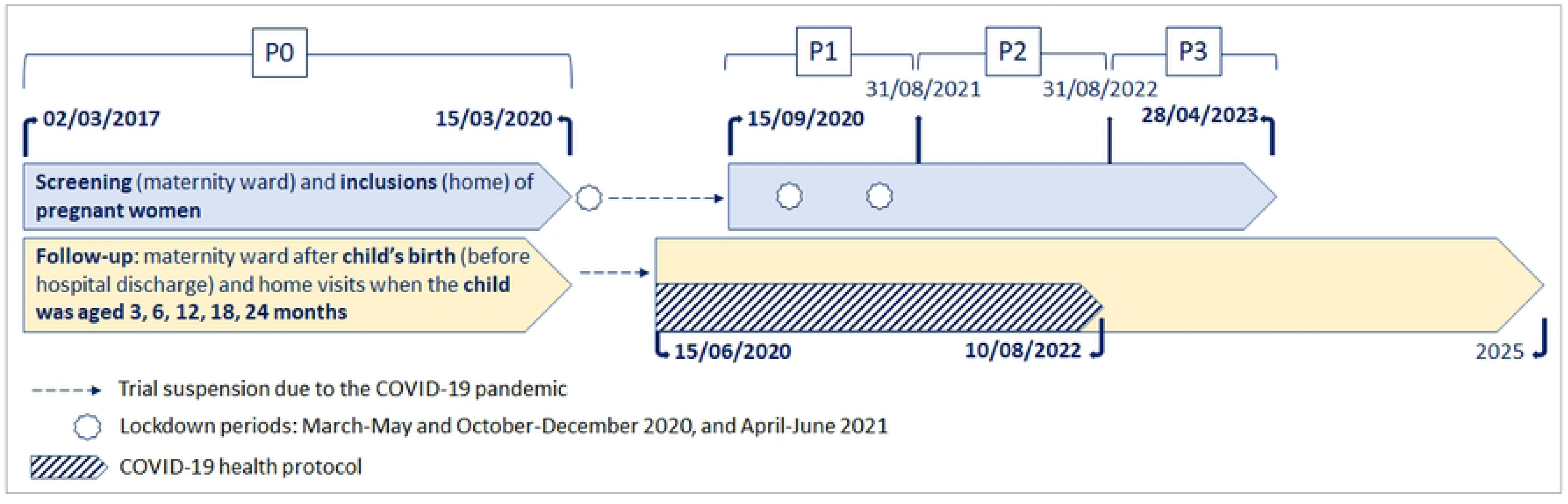
Timeline of ECAIL trial implementation in the Lille investigation area. The ECAIL trial.

While waiting for their prenatal appointment, pregnant women were given an eligibility questionnaire (**Supplementary document 1**) by reception staff and completed it, covering all inclusion and exclusion criteria (**Table 1**). Healthcare providers (HCPs; e.g., midwives, nurses, or obstetricians) used this questionnaire to determine eligibility for the ECAIL trial. When a woman was deemed eligible, the HCP was expected, as part of the screening protocol, to contact a dietitian from the ECAIL implementation team by telephone. The dietitian then provided the woman and her partner (when present) with information about the study in person, immediately after the HCP appointment. Within the following week, the same dietitian contacted the eligible woman by telephone to assess her willingness to participate. If she agreed, an appointment was scheduled at her home (or another location of her choice) for the baseline visit during the third trimester of pregnancy. If the eligible woman declined participation, the dietitian recorded her reasons on the eligibility questionnaire.

**Table 1.**
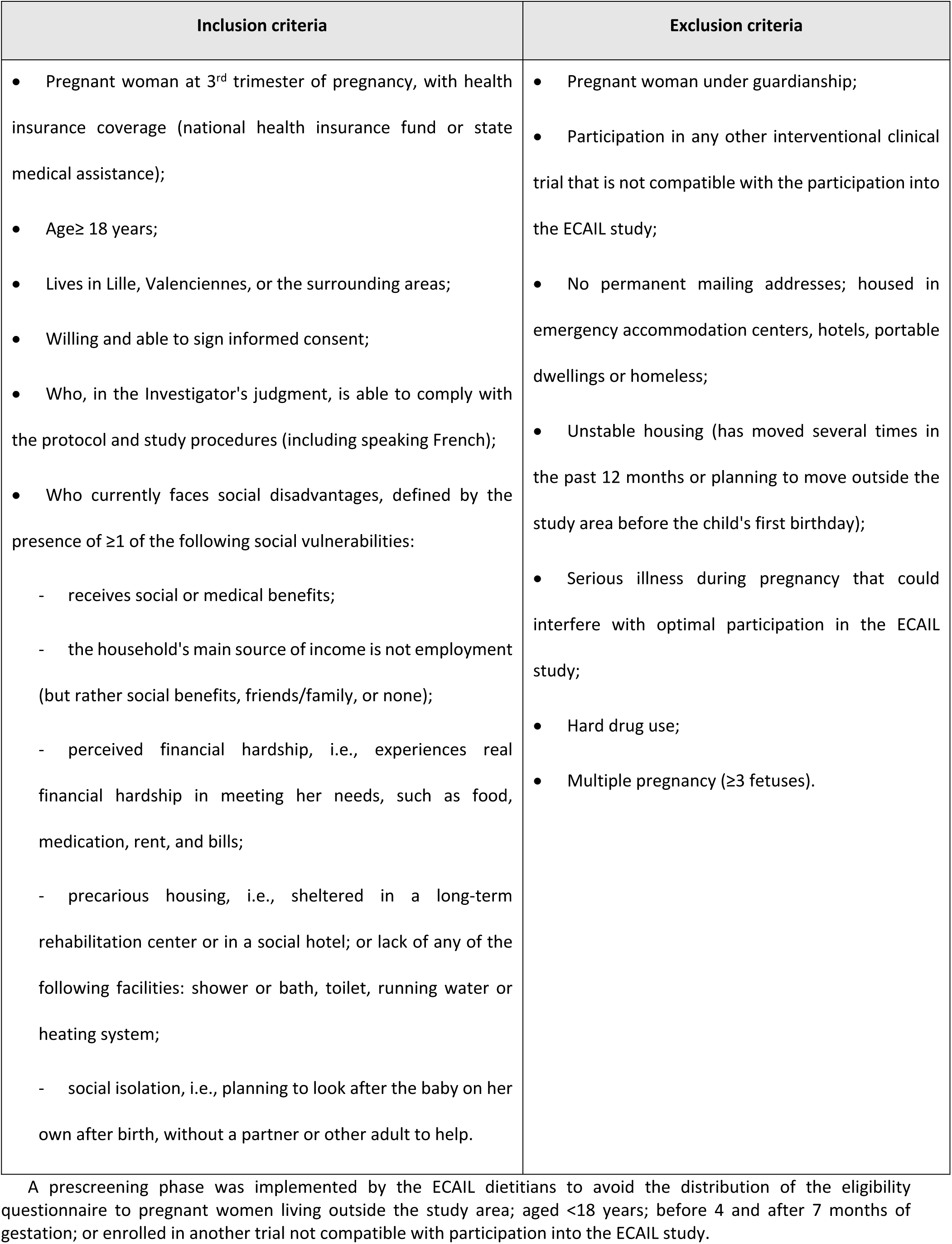
Inclusion and exclusion criteria of the ECAIL study, which the eligibility questionnaire screened for. The ECAIL trial.

When the HCP failed to call an ECAIL dietitian, he or she transmitted the questionnaire at a later time, and a dietitian then contacted the woman by telephone to provide study information. Women were then offered a one-week reflection period, followed by an additional call to discuss participation. Importantly, the provision of study information face to face by dietitians, rather than exclusively by telephone, served as an indirect indicator of HCP engagement with the screening procedure. Moreover, as part of the co-creation process, the eligibility questionnaire was co-designed, pilot-tested, and iteratively refined with HCPs, and monthly coordination meetings were used to identify barriers and collectively develop solutions to support implementation.

During the baseline home visit, the mother-to-be provided written consent for the participation of the mother-child dyad, randomization was performed, and the inclusion process was formalized. Subsequently, participating families were followed up by dietitians, first at the maternity ward shortly after delivery, and then through five additional home visits until the child’s second birthday, that is, at 3, 6, 12, 18, and 24 months.

Due to the COVID-19 pandemic, screening, inclusions, and home visits were suspended during the first national lockdown, which started on March 15, 2020 (Fig 1), when 140 families were actively under follow-up in the study. To enable the safe continuation of the intervention, implementation adaptations were introduced upon resumption. Home visits (June 2020) and screening/inclusions (September 2020) resumed under an adapted delivery protocol. Dietitians contacted participants by telephone before each home visit to administer a questionnaire assessing COVID-19 symptoms within the previous three weeks and recent contact with a confirmed case. Dietitians had the option to consult a physician to decide whether to proceed with the visit or postpone it for two weeks. Before the pandemic, dietitians typically sent a text message prior to visits without requiring participant confirmation; this pre-visit telephone contact was introduced after resumption. During home visits, dietitians were required to comply with protective measures, including wearing masks, practicing hand hygiene, and maintaining social distancing, and participants were requested to do the same. These adaptations allowed the intervention to be delivered while maintaining feasibility and in-person contact with families.

### Data collection and measurements

#### Time periods

We defined two periods, namely prepandemic (March 2017 to March 2020; P0) and post-study resumption (September 2020 through April 2023). To capture temporal variation in implementation conditions, the post-resumption period was segmented into three periods of approximately one year (September-August) to reflect post-resumption yearly cycles; the final period was shorter due to the end of screening in April 2023 (Fig 1; P1, P2, and P3).

#### Indicators of implementation

Drawing on the RE-AIM framework, the following indicators were selected: eligibility and participation as indicators of *Reach*; delivery modalities as indicators of *Implementation* and *Adoption* (as they reflect HCP engagement in screening and referral); and follow-up completion and adaptations as indicators of *Implementation* (19, 20).

### Eligibility

The eligibility rate was calculated as the number of eligible pregnant women (according to the responses collected from the eligibility questionnaires) divided by the number of eligibility questionnaires completed by pregnant women and returned to the ECAIL dietitians. As a proxy for implementation processes and provider engagement, the mode of study information delivery by dietitians to eligible women was categorized as either in person or by telephone.

### Participation

The participation rate was calculated as the number of actual inclusions in the ECAIL trial divided by the number of eligible pregnant women. Non-participation was classified into four categories: women who were unreachable; screening failures (i.e., cases in which inclusion could not be completed, regardless of the women’s decision, due to timing constraints, pregnancy outcomes, or unforeseen exclusion criteria); refusals without stated reasons; and refusals with one or more reasons, recorded verbatim by ECAIL dietitians.

### Follow-up

Based on the theoretical number of follow-up visits expected to take place from trial commencement to the end of the screening in April 2023, we calculated the rate of skipped visits. We also calculated the rate of failed visits, defined as cases in which the dietitian found nobody at the participant’s home, expressed as the number of failed visits divided by the number of trips made by dietitians.

#### Indicators of social vulnerability

Six binary variables (yes/no) were defined using information available in the eligibility questionnaire: receipt of social or medical benefits; household main income not derived from employment; perceived financial hardship; social isolation; precarious housing; and experiencing ≥3 of these social vulnerabilities. The latter indicator was defined as the sum of the first five social vulnerability variables, with “yes” coded as 1 and “no” as 0.

#### Focus group

A researcher in psychosocial sciences (DP) conducted an in-person focus group in May 2023 at the Lille University Hospital with the complete ECAIL implementation team, comprising six dietitians (including the coordinator), to explore the impact of the COVID-19 pandemic on trial implementation. Written informed consent was obtained from each dietitian.

Several considerations guided the use of a focus group (26, 27). First, the COVID-19 pandemic substantially disrupted usual implementation conditions, warranting an in-depth qualitative assessment alongside quantitative indicators. Second, dietitians were identified a priori as the most relevant stakeholders for this analysis, given their central role and sustained engagement with study participants throughout the intervention. Third, as the dietitians shared a common implementation context, a focus group facilitated the collection of shared perspectives, experiences, and adaptations. This method was particularly suited to capturing local contextual influences on intervention delivery.

The ECAIL RCT has approval from the committee for protection of persons engaged in research (*Comité de Protection des Personnes,* CPP, 24/07/2014), the national advisory committee on information processing in health research (*Comité consultatif sur le traitement de l’information en matière de recherche,* CCTIRS, 04/09/2014), the national data protection authority (*Commission nationale de l’informatique et des libertés*, CNIL, 15/06/2016). The protocol for the qualitative component was approved by the CPP the 28/06/2021. Written and signed consent were obtained from participants.

### Analysis

#### Quantitative analysis

In addition to the eligibility rate, we compared the following implementation-related indicators between the prepandemic and post-resumption periods:

- among eligible women: participation rate, distribution of social vulnerability indicators, and mode of delivery of study information (in person or by telephone);
- among nonparticipating women: distribution of the main reasons for not participating;
- among participating women: rates of both skipped and failed follow-up visits;
- among ineligible women: proportion presenting at least one indicator of social vulnerability, along with the distribution of their social vulnerabilities and exclusion criteria.

Two-sided χ2 tests were used for categorical variables.

All analyses were conducted using RStudio (2022) (RStudio: Integrated Development Environment for R. RStudio, PBC, Boston, MA).

#### Qualitative analysis

DP manually transcribed the focus group verbatim and imported it into NVivo software. A thematic content analysis (28) was then conducted to identify key themes related to implementation context, disruptions, adaptations, and perceived mechanisms influencing reach and engagement. Relevant quotations were assigned to corresponding themes and subthemes that reflected dietitians’ shared experiences of trial implementation during and after the pandemic.

## Results

Results are presented across three RE-AIM–relevant dimensions in disrupted contextual conditions, namely *Reach*, *Adoption*, and *Implementation*.

### Descriptive findings from the quantitative component

#### Eligibility for the ECAIL trial

In all, 5,744 pregnant women completed the eligibility questionnaire between March 2017 and April 2023 at the Lille University Hospital maternity ward; 1,804 were deemed eligible for the ECAIL trial. Among the women screened, 29.7% (n=955) were eligible in the prepandemic period and 33.6% (n=849) post-resumption (*P*=0.002). The eligibility rate remained consistently higher throughout the post-resumption observation period (P1–P3) (**Fig 2**).

**Fig 2.**
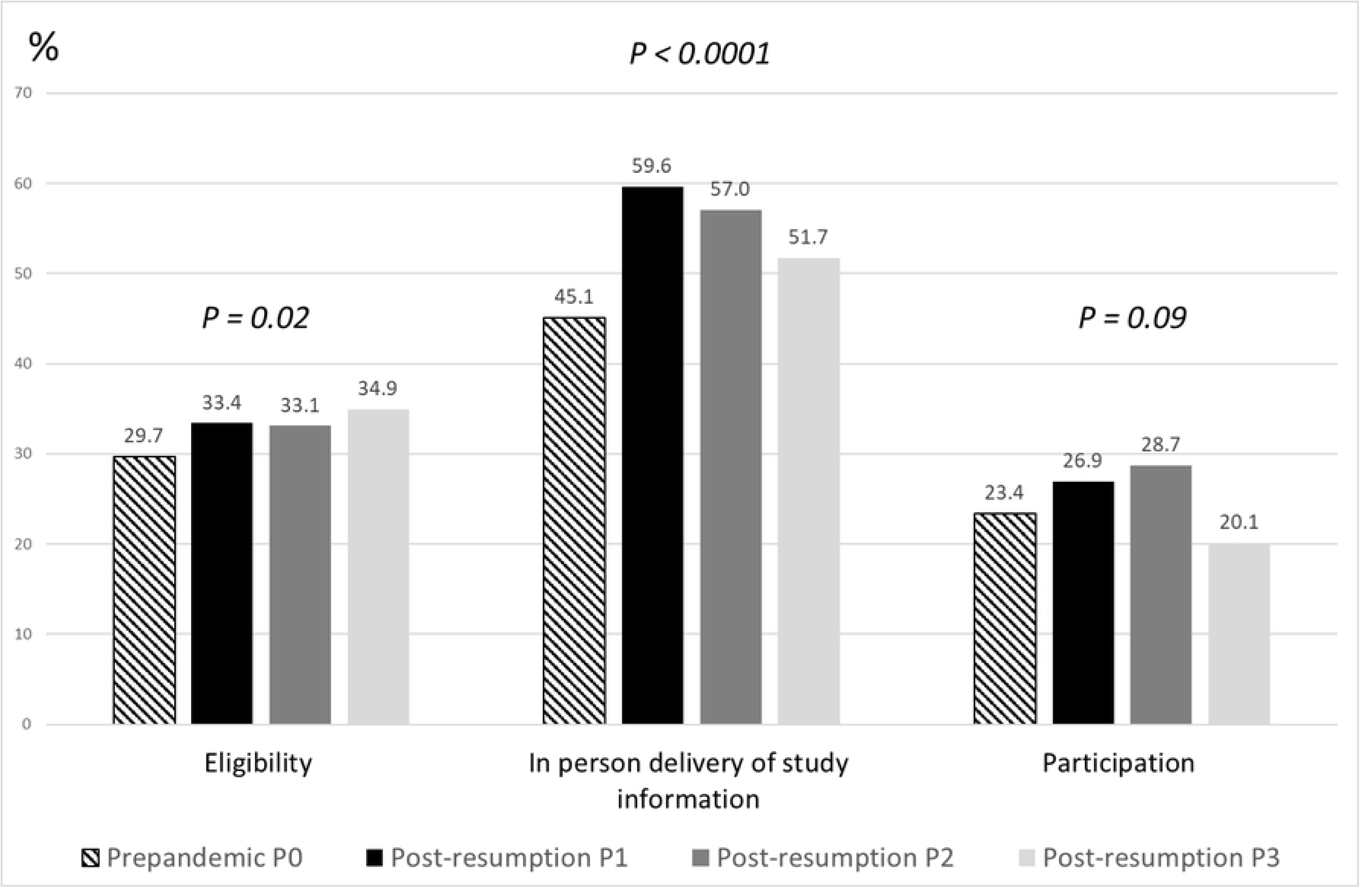
Rates of eligibility for the ECAIL trial (n=1,804), and rates of in-person delivery of study information (n=915) and participation (n=443) among eligible women according to the four study periods; prepandemic, P0: March 2017 to March 2020; post-resumption, P1: September 2020 to August 2021; P2: September 2021 to August 2022; P3: September 2022 to April 2023. The ECAIL trial, Lille investigation area.

Overall, no significant differences were observed in the distribution of social vulnerability indicators among eligible women between the prepandemic and post-resumption periods (**Table 2**): 78.3% received social or medical benefits; employment was not the main source of household income for 58.7%; 24.4% experienced financial hardship; 14.7% reported social isolation; 6.0% lived in precarious housing; and 19.0% presented three or more of these social vulnerabilities. However, when examined across post-resumption subperiods, **Fig 3** shows that the proportion of eligible women for whom the main source of income in the household did not derive from employment was highest during P1 (64.8%), as was the receipt of social or medical benefits (82.8%). The proportion of eligible women perceiving financial hardship was highest during P3 (29.3%). Only 50.7% of eligible women received information on the study in person from the ECAIL dietitians, rather than by telephone, but this rate was higher post-resumption (57.0% compared with 45.1% before the pandemic, **Table 2**). **Fig 2** shows that this rate was highest during P1 (59.6%) and declined progressively to 51.7% in P3.

**Table 2.** Distribution of social vulnerabilities and study implementation indicators among eligible women by period (%). The ECAIL trial, Lille investigation area.

|  | March 2017<br>to Apr 2023<br>(n=1,804) | March 2017<br>to March<br>2020<br>(n=955) | Sept 2020<br>to Apr<br>2023<br>(n=849) | P-value |
| --- | --- | --- | --- | --- |
| **Social vulnerabilities** |  |  |  |  |
| Receives social or medical benefits | 78.3 | 77.2 | 79.5 | 0.23 |
| Household's main source of income is not employment | 58.7 | 56.8 | 60.9 | 0.07 |
| Perceived financial hardship | 24.4 | 25.8 | 23.0 | 0.17 |
| Social isolation | 14.7 | 14.3 | 15.1 | 0.66 |
| Precarious housing | 6.0 | 5.5 | 6.6 | 0.35 |
| ≥ 3 social vulnerabilities | 19.0 | 18.8 | 19.2 | 0.81 |
| **Study implementation indicators** |  |  |  |  |
| *Screening (n=1789)* |  |  |  |  |
| In-person delivery of study information | 50.7 | 45.1 | 57.0 | <0.0001 |
| *Follow-up (n=444)* |  |  |  |  |
| Theoretical number of visits expected | 2316 | 919 | 1397 |  |
| Skipped visits, % (n) | 2.0 (46) | 1.7 (16) | 2.1 (30) | 0.49 |
| Failed visits, % (n) | 3.0 (70) | 4.9 (47) | 1.7 (23) | <0.001 |

**Fig 3.**
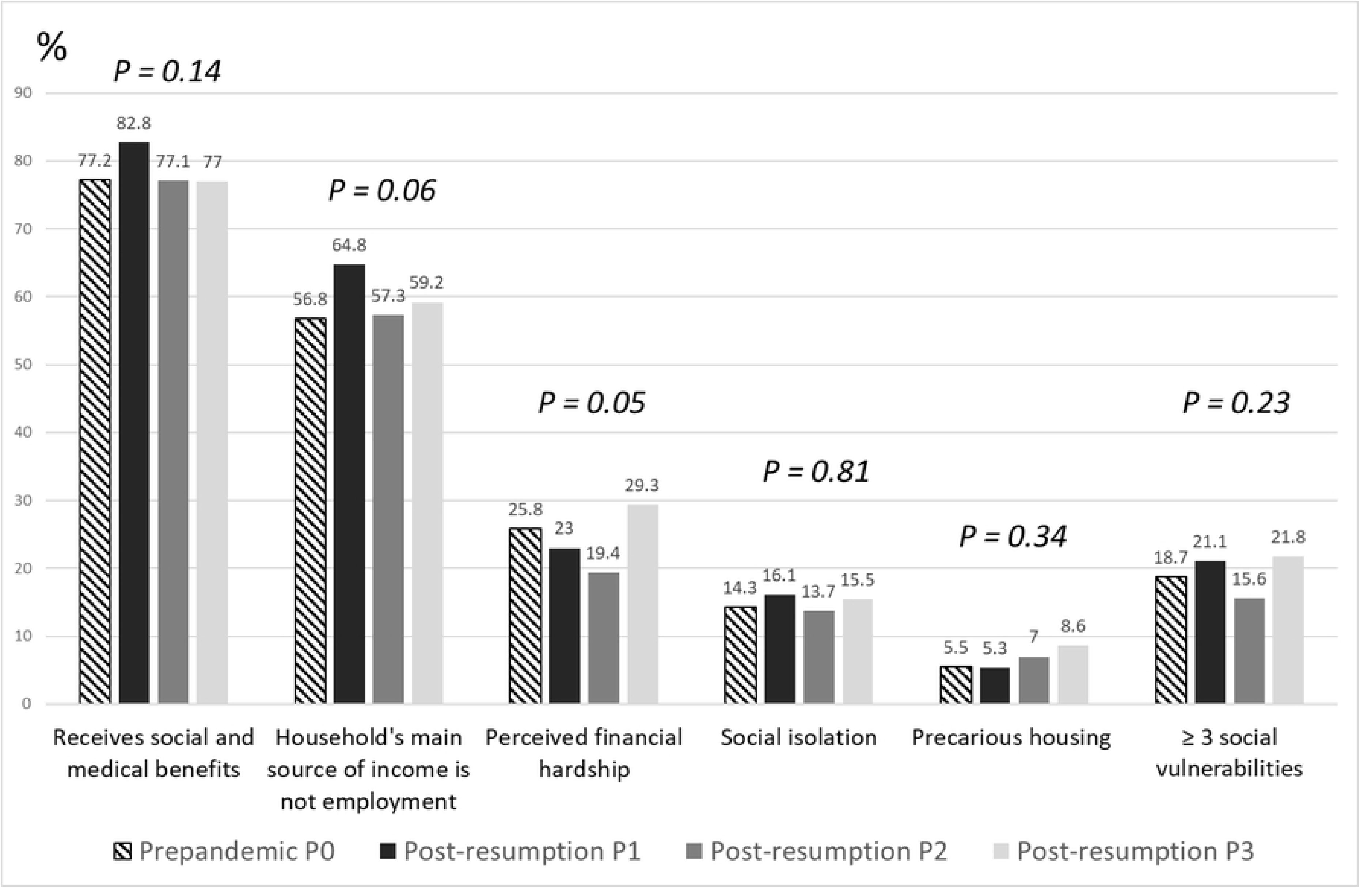
Distribution of social vulnerabilities among eligible women (n=1,804) according to the four study periods; prepandemic, P0: March 2017 to March 2020; post-resumption, P1: September 2020 to August 2021; P2: September 2021 to August 2022; P3: September 2022 to April 2023. The ECAIL trial, Lille investigation area.

#### Participation in the ECAIL trial

Overall, 24.6% of eligible women consented to participate and were included in the trial, with no difference in participation rates between prepandemic and post-resumption periods. However, participation rose slightly during periods P1 (26.9%) and P2 (28.7%), before declining in period P3 (20.1%), falling below the prepandemic level observed in P0 (23.4%) (**Fig 2**). The distribution of social vulnerability and implementation-related indicators differed between participating and nonparticipating women (**Table 3**). Across both periods, women who agreed to participate more frequently reported perceived financial hardship and were more often informed about the study in person compared with those who declined.

**Table 3.** Characteristics of eligible women by participation status and study period (prepandemic and post-resumption) (%). The ECAIL trial, Lille investigation area.

|  | March 2017 - March 2020 |  |  |  | Sept 2020 - Apr 2023 |  |  |
| --- | --- | --- | --- | --- | --- | --- | --- |
|  | n=955 |  |  |  | n=849 |  |  |
|  | Participation | Non-participation | P-value |  | Participation | Non-participation | P-value |
| **Social vulnerabilities** |  |  |  |  |  |  |  |
| Receives social or medical benefits | 74.9 | 77.9 | 0.35 |  | 77.5 | 80.2 | 0.38 |
| Household's main source of income is not employment | 57.8 | 56.4 | 0.71 |  | 57.7 | 62.0 | 0.25 |
| Perceived financial hardship | 37.2 | 22.3 | <0.0001 |  | 29.7 | 20.6 | 0.005 |
|  | n=955 |  |  |  | n=849 |  |  |
|  | Participation | Non-participation | P-value |  | Participation | Non-participation | P-value |
| Social isolation | 15.7 | 13.9 | 0.51 |  | 14.4 | 15.3 | 0.75 |
| Precarious housing | 7.2 | 5.1 | 0.23 |  | 3.2 | 7.8 | 0.02 |
| ≥ 3 social vulnerabilities | 24.7 | 16.9 | 0.01 |  | 17.1 | 19.9 | 0.36 |
| **Study implementation** |  |  |  |  |  |  |  |
| In-person delivery of study information | 54.3 | 42.3 | 0.002 |  | 64.9 | 54.2 | 0.006 |

#### Nonparticipation in the ECAIL trial, despite eligibility

There was no major difference in the overall pattern of nonparticipation between the prepandemic and post-resumption periods: a majority of eligible women either remained unreachable or declined participation without providing a reason, while 29% gave at least one reason for nonparticipation (**Fig 4**). Screening failures primarily occurred when pregnancies were too advanced or births took place before the scheduled baseline visit, with the latter more frequent post-resumption, reflecting the suspension of the inclusion process during the first lockdown (**Fig 5**). Among women who provided reasons for nonparticipation, time and availability constraints were reported more frequently post-resumption, whereas partner or relative refusal and having older children were less common (**Fig 6**).

**Fig 4.**
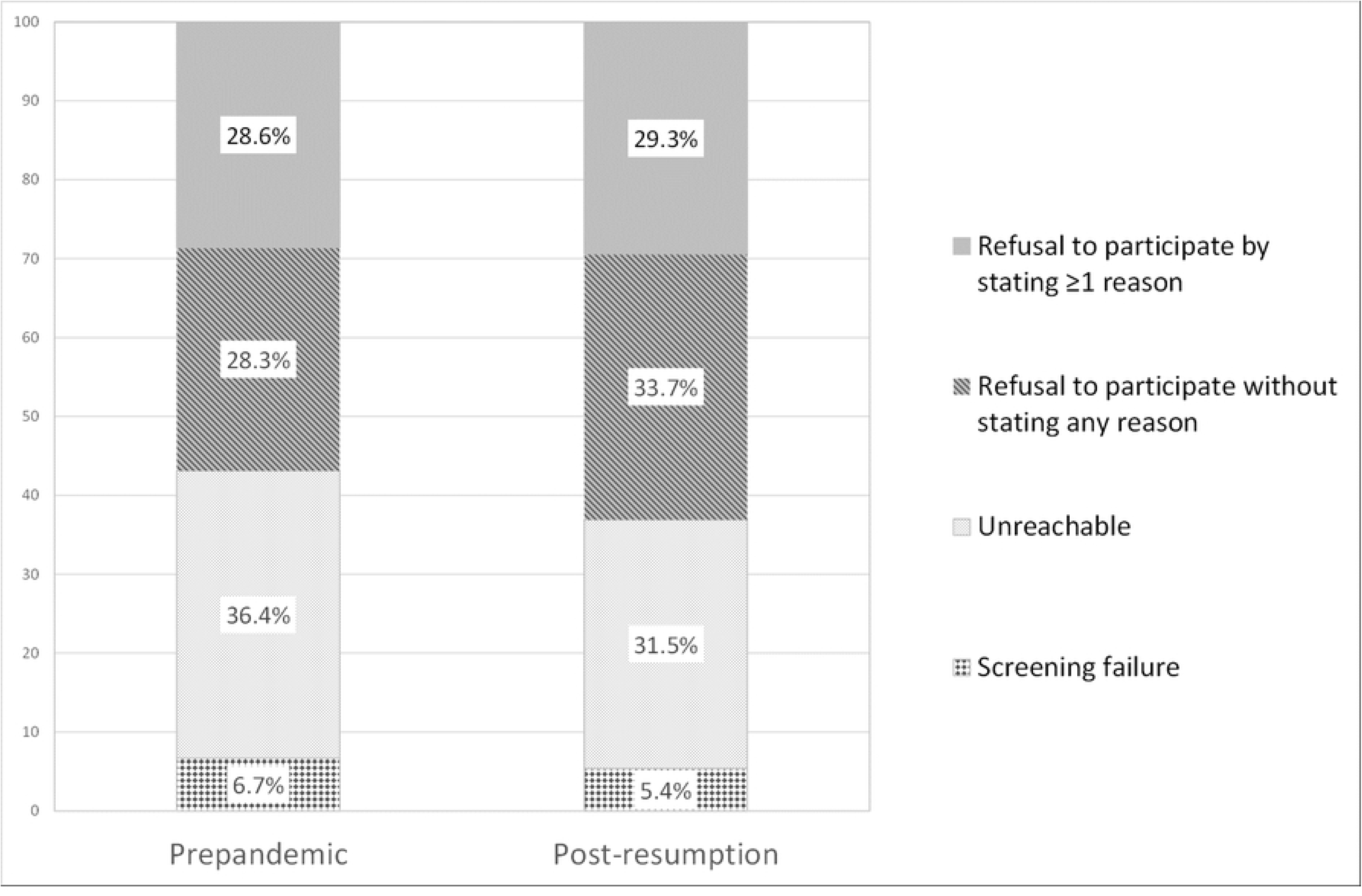
Distribution of the four main reasons for nonparticipation in the study among eligible nonparticipating women (n=1345) by period (prepandemic and post-resumption). The ECAIL trial, Lille investigation area.

**Fig 5.**
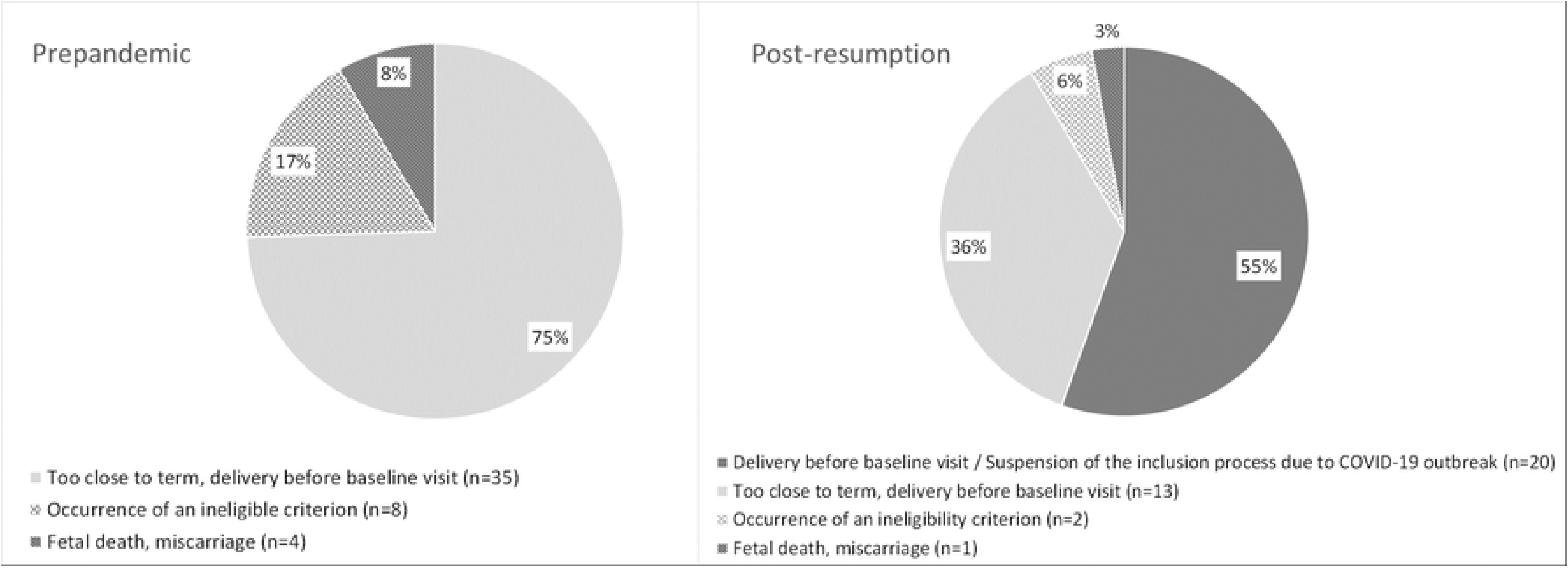
Characterization of screening failures by study period (prepandemic and post-resumption). The ECAIL trial, Lille investigation area.

**Fig 6.**
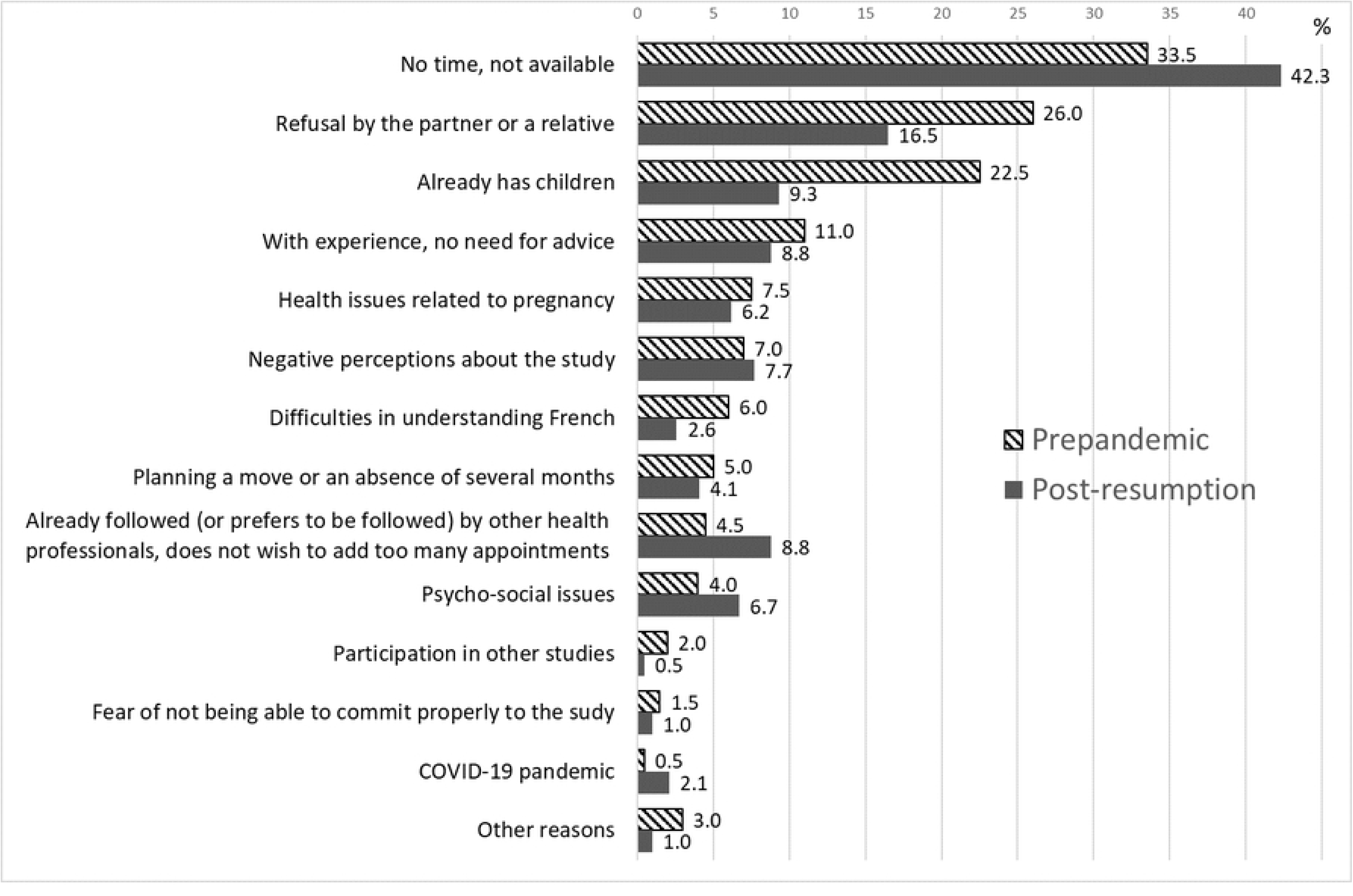
Distribution of the reasons for nonparticipation stated by 396 women by study period (prepandemic and post-resumption). The ECAIL trial, Lille investigation area. *Negative perceptions about the study* include “Too long”, “Too constraining”, “Not useful”; *Psycho-social issues* include “Personal reasons”, “Overwhelmed”, “Too many problems to deal with”, “Health issues with a child or another family member”, “Too instable”, “Inability to plan ahead”; *Participation in other studies* includes in the past or currently.

#### Ineligibility for the ECAIL trial despite social disadvantage

Among ineligible women, 19.1% (n=746) were identified as experiencing at least one social vulnerability (according to the eligibility questionnaire) but also met at least one exclusion criterion. Their profile of social vulnerability was overall more severe than that of eligible women, particularly regarding unemployment, perceived financial hardship, precarious housing, and accumulation of ≥3 of these social vulnerabilities criteria (**Table 4**). Furthermore, among socially disadvantaged but ineligible women, the proportion experiencing social isolation during their pregnancy increased from 15.0% prepandemic to 21.5% post-resumption, while the proportion reporting precarious housing rose from 11.7% to 19.8%. Likewise, housing instability worsened across periods among socially vulnerable women meeting exclusion criteria: the proportion who had moved several times in the previous 12 months increased from 16.6% prepandemic to 25.3% post-resumption, and the proportion living in emergency accommodation, portable dwellings, or who were experiencing homelessness increased from 7.1% to 15.4%.

**Table 4.** Distribution of inclusion and exclusion criteria among women facing social disadvantage ineligible for the ECAIL study by study period (%). The ECAIL trial, Lille investigation area.

|  | March 2017 to<br>Apr 2023<br>N=746 |  | March 2017 to<br>March 2020<br>n=453 | Sept 2020 to<br>Apr 2023<br>n=293 | P-value |
| --- | --- | --- | --- | --- | --- |
| **Inclusion criteria: social vulnerabilities** |  |  |  |  |  |
| Receives social or medical benefits | 75.1 |  | 77.0 | 72.0 | 0.12 |
| Household's main source of income is not employment | 65.1 |  | 65.3 | 64.8 | 0.89 |
| Perceived financial hardship | 31.0 |  | 31.1 | 30.7 | 0.91 |
| Social isolation | 17.6 |  | 15.0 | 21.5 | 0.02 |
| Precarious housing | 14.9 |  | 11.7 | 19.8 | 0.002 |
| ≥ 3 social vulnerabilities | 27.1 |  | 25.8 | 29.0 | 0.34 |
| **Exclusion criteria** |  |  |  |  |  |
| Planning to move outside the study area before the child's first birthday | 30.0 |  | 34.4 | 23.2 | 0.001 |
| Moved several times in the past 12 months | 20.0 |  | 16.6 | 25.3 | 0.004 |
| Sheltered in emergency centers, portable dwellings or homeless | 10.3 |  | 7.1 | 15.4 | <0.001 |
| Suboptimal understanding/speaking of French | 20.6 |  | 17.9 | 24.9 | 0.02 |
| Serious illness during pregnancy | 18.2 |  | 17.9 | 18.8 | 0.15 |
| Under guardianship | 5.0 |  | 3.8 | 6.8 | 0.059 |
| Multiple pregnancy (≥3 fetuses) | 1.1 |  | 1.5 | 0.3 | 0.16 |
| Hard drug use | 2.4 |  | 1.8 | 3.4 | 0.15 |

#### Follow-up

From March 2017 to April 2023, only 2% (n=46) of the 2,316 expected follow-up visits were skipped, with no difference between prepandemic and post-resumption periods (Table 2). The rate of failed visits (the dietitian made the trip but the participant was not present) was, however, lower after the study resumed (1.7%) than before the COVID-19 outbreak (4.9%).

### Findings from the qualitative component

The focus group, which lasted 81 minutes, explored how COVID-19 affected home visit delivery, strategies to maintain relationships with participating families, the pivotal moment of the resumption of home visits, and the acceptability of the adapted health protocol for both dietitians and families. **Table 5** presents the key themes and illustrative quotations. During the first lockdown, dietitians expressed uncertainty and concern about whether, when, and how the study would resume. Despite obstacles, they were resolute in implementing the study through home visits, arguing that the remote solutions developed during the lockdown did not fit the trial’s specific features. Their point of view was important in designing the subsequent adaptations, as they took part in discussions surrounding implementation under new constraints, and they anticipated resumption by maintaining contact with participants through telephone calls and newsletters. These adaptations appeared to have strengthened engagement and commitment when home visits became possible again. Both dietitians and participants (as reported by dietitians) expressed gladness at the resumption of the trial. The ECAIL study was prioritized by Lille University Hospital over other trials because of the social vulnerability of its participants. Dietitians reported that mothers described these visits as helping to alleviate social isolation, which had intensified during successive lockdowns. Dietitians did not perceive mask-wearing during hospital screenings or home visits as a barrier to women’s participation or to building rapport during follow-up visits, given the widespread use of masks at the time. Yet they allowed themselves to relax mask use during home visits, with mothers’ authorization, in cases where it hindered interactions with the child. Finally, the pre-visit COVID-19 risk assessment questionnaire was perceived as an effective implementation adaptation, helping to reduce failed visits and save time.

**Table 5.** Qualitative themes retrieved from the group interview and mapped onto RE-AIM dimensions, with illustrative quotations.

| RE-AIM dimension | Themes | Sub-themes | Illustrative quotations |
| --- | --- | --- | --- |
| Implementation | Theme 1.<br>Maintaining contact with participants while visits were suspended | Proactive relational continuity | *"We stopped but we still kept contact, in any case I kept the link by phone with the ladies, especially when there were the visits that arrived, those of the three months, during that period, in any case I called all the ladies to explain to them that for the moment the ECAIL visits were stopped and that we would keep them informed, we even made a little note ... the little newsletter with a little message to encourage them with our photos, it was still important to keep this link even if we didn't really know what to say to them, but still to say that we were there and that they could contact us if necessary, that we were available by phone." - Dietitian 1* |
| Implementation |  | Maintaining link facilitated commitment | *"That was good, and after, contacting them again to restart the visits, adherence immediate." - Dietitian 1* |
| Adoption | Theme 2.<br>Resuming home visits | Uncertainty regarding the resumption of home visits | *"In any case, for me, it was a fear of telling myself how we were going to return to people afterward? Can we go back? Because it was so unclear at the beginning, everything was at a standstill ... And therefore to tell ourselves how we are going to find the ladies? Do we leave them like this? [...] Will we be able to resume? And if we resume will it always be, will it be the same?" - Dietitian 1* |
| Reach |  | Participants' happiness at the resumption of home visits | *"The ladies were glad to see people again, and so, I remember that the schedule was full of visits because they were glad ... the ladies were really, really happy to have someone come to the house because it was still the lockdown, so someone coming to the house to chat ... glad to have a social link with someone from outside again, not having their family locked inside their apartment." - Dietitian 2* |
| Adoption |  | Dietitians' engagement | *"I was also glad to resume the visits." - Dietitian 2* |
| Implementation |  | Necessity of maintaining home-visits as a core feature of the intervention | *"For us, with ECAIL visits, video calls aren't even an option; we can't do the visit via video (Dietitian 1). Not even phone calls? (DP). No (Dietitian 5). We can't do a visit over the phone; it's not possible. All the tools that were put in place as we went along just didn't suit us (Dietitian 1)"* |
| Implementation |  | Longer visits after resumption | *"However, clearly, the visits, at least in my recollection, I'm not sure if it's biased by the fact that it's dated, but the visits lasted a long time." - Dietitian 2* |
| Adoption |  | Prioritization of the ECAIL study resumption among other trials | *"And many protocols for healthy volunteers didn't resume, didn't resume at that time. They made a real exception for the ECAIL study because of its specific audience, and we measured that there were more benefits in going back to people than waiting further to avoid the risk of contamination." - Dietitian 5* |
| Implementation | Theme 3.<br>Adaptation of protocol | Health protocol as a condition for visit resumption | *"It's true that we were allowed to resume visits in June only if we implemented this health protocol ... we had to implement this protocol to protect both the families and the dietitians." - Dietitian 5* |
| Implementation |  | Acceptability of masks for participants | *"We saw the ladies as part of the continuation of their prenatal consultation ... it was something from everyday life [the mask] it was mandatory in public places, in public transportation ... so it became customary, actually. So even more at the hospital, the place where it is necessary to wear the mask. So, as a result, I think that it was logical, well, it was the most logical place, and I don't think that it posed a problem for them, that it was a barrier." - Dietitian 1*<br>*"In fact, it was an everyday thing, the mask eventually became part of daily life, so it wasn't something that presented a barrier." - Dietitian 1* |
| Implementation |  | Acceptability of masks for children | *"I can see that the mask is a barrier for the children, so I take it off in those situations; if I see that it's too difficult for the child to see me with a mask on, I ask the lady if that's possible"* - Dietician 6 |
|  |  | Pre-visit COVID-19 risk assessment questionnaire: reduction of missed visits and time-saving | *"It's true that we had this reflection, well, we arrived directly with the COVID questionnaire, there were many fewer trips with no one home, for nothing."* - Dietitian 3 |
|  |  |  | *"So, this allowed for efficiency, and, well, it had unforeseen positive consequences, in the sense that there were fewer unnecessary trips for nothing, less time wasted."* - Dietitian 5 |

## Discussion

This interdisciplinary study of an ongoing public health intervention among socially disadvantaged families provides implementation-relevant insights into the effects of the COVID-19 pandemic and the subsequent cost-of-living crisis. Interpreted through a RE-AIM lens (19, 20), our findings indicate that these contextual shocks expanded the pool of eligible individuals, enhancing the intervention’s relevance (*Reach*), without compromising provider *Adoption* or *Implementation*, and possibly reinforcing engagement during key phases of delivery. Quantitative and qualitative findings converged, showing shifts in eligibility and participation profiles alongside sustained involvement of both implementers and participants, resulting in a stable rhythm of home visits after resumption, with some parents eager to re-establish social contact. These health and economic shocks also affected socially disadvantaged families beyond those eligible for the ECAIL trial, exacerbating existing social and material vulnerabilities. Taken together, these findings point to robust implementation and short-term maintenance despite major contextual disruptions and inform the design of more resilient interventions.

### Eligibility, ineligibility, and social vulnerabilities

Hauts-de-France is one of the regions with the highest poverty rates in France. According to Insee, in 2020, 17.2% of its population live below the poverty line (29), compared with a national average of 13.6% (30). Of the region’s four districts, *Nord* (where Lille is located) has the highest poverty rate (18.4%) (29), reflecting the sharp economic decline of the 1980s following the closure of coal mines and the collapse of the textile and steel industries (31). Its working population has long included a higher proportion of blue-collar and hourly workers than the national average (32, 33), and unemployment has remained relatively high (34). Only 25.9% of the Hauts-de-France population held a postsecondary degree in 2020 (35), compared with 31.6% nationally (33). Furthermore, single parents headed 17% of families, 85% of whom are single mothers (36). Compared with two-parent households, single mothers participate less frequently in the labor force and, when employed, are more often in lower-paid occupations; over two-thirds of them are hourly or manual workers (37). These structural characteristics likely explain the consistently high eligibility rate observed in the ECAIL trial, even before the pandemic.

Against this already disadvantaged socioeconomic backdrop, the COVID-19 pandemic further exacerbated existing inequalities. Overall, the pandemic hit low-income households hardest, as they were more likely to work in heavily affected sectors, such as catering, transportation, blue-collar occupations, and informal work, and less likely to hold long-term contracts and positions compatible with teleworking (38). As a result, household living standards declined. Labor-market disruptions, including those affecting lower middle-class households, likely contributed to the higher post-resumption eligibility rate, consistent with the increased proportion of women reporting that employment was not their household’s main source of income during P1. Likewise, at the national level, the poverty rate increased from 13.6% in 2020 to 15.4% in 2023 (30).

National administrative data showed sharp increases in benefit receipt early in the pandemic (39). Consistently, we observed a higher proportion of ECAIL-eligible women receiving social or medical benefits during P1, which may help explain the concurrent decrease in perceived financial difficulties. This apparent buffering effect may also reflect short-term changes in household consumption during lockdowns, when reduced spending opportunities enabled some families to accumulate savings. In 2020, household consumption expenditure fell by 7.1%, particularly for accommodation, catering, transport, leisure, and culture (40).

The eligibility rate nonetheless remained relatively high during P2 and P3, likely reflecting persistent macro-economic pressures, including rising energy and food prices. Inflation disproportionately affected low-income households, due to the composition of their expenses and the poverty penalty described earlier. Though employment rates partially recovered as health restrictions were lifted, the withdrawal of exceptional support measures may have contributed to the increased reporting of financial difficulties during P3. More broadly, the cost-of-living crisis further compounded the challenges faced by a population already weakened by the health crisis. According to the 17^th^ *Ipsos/Secours Populaire Poverty Barometer*, difficulties in meeting everyday expenses intensified further in 2023, particularly regarding children’s needs, health care, energy, and food (41). These trends were mirrored by a marked increase in food insecurity in France, with the proportion of respondents unable to consume fresh fruit and vegetables daily rising from 27% in 2018 to 43% in 2023.

This worsening socioeconomic context is also reflected in our analysis of women ineligible for the ECAIL trial despite experiencing social vulnerability. These women faced increasing housing precarity and instability, as well as heightened social isolation. These findings are consistent with those reported by Baillieul and Braun (2021) (42), who found that in the Hauts-de-France region, 10% of households with the lowest incomes experienced a 32% decline in their standard of living.

Taken together, these results underscore how multidimensional crises such as the COVID-19 pandemic can intensify pre-existing vulnerabilities, highlighting the need for resilient intervention designs.

### Participation

Participation in the ECAIL trial increased slightly during the post-resumption P1 and P2 subperiods. From an *Implementation* perspective, it confirms the dieticians’ impression that screening interviews were unaffected by the protective measures such as masks. From an *Adoption* perspective, this was likely associated with a higher frequency of face-to-face delivery of study information at the maternity ward, rather than telephone contact. Direct interactions between dietitians and women (and their partners, when present) appeared more effective in initiating engagement than delayed, impersonal phone calls, although this depended on HCPs contacting dietitians during prenatal appointments. The prioritization of the ECAIL trial likely created favorable conditions for its implementation, reflecting an equity-oriented approach. In this context, regular meetings between the ECAIL team and HCPs likely supported sustained and synergistic collaboration despite pandemic-related pressures on hospital services. Together, quantitative and qualitative findings converged to suggest strengthened coordination between dietitians and HCPs during the early post-resumption period (P1). However, HCP engagement appeared to decline slightly during period P3, in parallel with the decrease in participation. This may reflect increased workload following the resumption of other clinical trials, compounded by persistent disruptions and time constraints affecting hospital services. The slight decrease in participation during P3 may also reflect the cumulative burden of successive crises, which likely reduced women’s availability and willingness to participate, as suggested by the more frequent reporting of time constraints and unavailability among those who declined participation post-resumption. These findings highlight the prolonged and evolving impact of crises, underscoring the importance of considering temporal dynamics in their effects.

### Follow-up

Across both qualitative and quantitative components, there was clear evidence that dietitians and families were eagerly awaiting the resumption of visits. During the first lockdown, dietitians maintained regular contact with participants. This continuity of contact emerged as a key implementation mechanism for sustaining family engagement, as reflected in the very low rate of skipped home visits. The implementation of COVID-19 -specific health protocols facilitated preliminary exchanges, reduced potential apprehension among participants, and contributed to fewer missed appointments.

These observations have significant methodological implications and highlight the value of ongoing relational work between follow-up visits. This requires additional human resources and research time, particularly when working with socially disadvantaged populations, who are known to be “hard-to-survey” (21, 22). However, this investment appears cost-effective, as it saves time in the long term, improves adherence to the study protocol, fosters participant commitment, and enhances data quality by limiting missed visits. More broadly, our findings reinforce the relevance of home-based delivery for reaching and engaging populations facing social adversity, who often have lower utilization of health care and community services and experience greater social isolation (43, 44), while facilitating one-on-one interactions that foster trust between providers and mothers and enable more personalized and context-sensitive interventions (18).

### Perspectives

This study examined how the distribution of social vulnerability criteria differed between eligible and ineligible women, as well as between participants and nonparticipants, an aspect that remains rarely explored in implementation research (44). It also illustrates how such data can be collected and analyzed to evaluate intervention reach (45). These findings provide important insights for interpreting the external validity of the trial and the potential generalizability of future effectiveness results. Additionally, qualitative interviews are currently being conducted with participating mothers to explore their motivations, contextual constraints, and perceptions of participation. These complementary data will contribute to a more comprehensive process evaluation of this complex public health intervention (46–49).

### Limitations and strengths

One limitation of this study is the timing of the group interview, conducted two years after the last lockdown, which may have introduced a recall bias. Nonetheless, the vividness of the dietitians’ recollections underscores the lasting impact of this period and the relevance of capturing these experiences retrospectively. While effectiveness outcomes are not examined here, this focus is consistent with the RE-AIM framework’s emphasis on disentangling *reach*, *adoption*, and *implementation* processes as necessary precursors to effectiveness and sustainability (19). A major strength lies in the triangulation of quantitative and qualitative methods (27, 44, 47), which enabled a comprehensive understanding of how successive crises shaped both the characteristics of the target population and the implementation processes of the intervention. Importantly, the inclusion of dietitians’ perspectives represents a further strength, as the views of frontline staff, particularly those directly involved in participant interaction and data collection, are still relatively rarely captured in implementation research. The study’s focus on pregnant women experiencing social vulnerabilities, a heterogeneous population that remains largely underrepresented in epidemiological research, provides new knowledge about the specific barriers to intervention delivery and engagement in a context of worsening poverty. The adjustments made after the onset of the COVID-19 pandemic illustrate the flexibility of this complex interventional research in responding to unforeseen events and adapting implementation to enable study continuation (46, 48). They also suggest that home visits and one-to-one interactions may strengthen participation and commitment, even when face-to-face contact is constrained.

## Conclusion

The social impact of both the pandemic and the cost-of-living crisis, often described as the “second health emergency” (5), is reflected in the ECAIL trial by a higher eligibility rate among socially disadvantaged families, while the participation rate remained stable. These findings provide important insights into how crises affect the ability of vulnerable families to engage in programs designed to mitigate social disadvantage. In the case of ECAIL, the health crisis did not undermine stakeholder or participant commitment to this co-created intervention. On the contrary, the collaborative foundations of the project appear to have supported sustained engagement by HCPs despite increased workload and organizational strain within hospitals. HCPs reported remaining committed to seeing the project through to completion, recognizing its social and public health importance. Moreover, home visits and telephone calls between follow-ups were perceived as effective implementation strategies, not only for reaching families and supporting adherence, but also for mitigating social isolation among participants. Overall, these findings suggest that interventions co-designed with stakeholders and target populations, and supported by appropriate implementation strategies, can remain resilient even in the face of major external disruptions, an insight that is particularly relevant in a context of rising poverty and ongoing geopolitical and economic instability worldwide.

## Data Availability

All data supporting the findings of this study are included in the present article or the supplemental material.

## Abbreviations

ECAIL: prEgnanCy and eArly Childhood nutrItion triaL;
HCP: healthcare provider;
Insee: National Institute of Statistics and Economic Studies;
Inserm: National institute for health and medical research;
RE-AIM: Reach, Effectiveness, Adoption, Implementation and Maintenance.

## Acknowledgments

The authors thank all stakeholders involved in the ECAIL study since 2012 for its co-design and since 2017 for its implementation, i.e., members from the Malin program nonprofit association involved in the co-creation of the ECAIL trial and/or the training of ECAIL dietitians, namely: Solène Bonhoure, Marie-Cécile Bret, Louise Jolly, Nelly Perez, Marie Polycarpe, Julie Simon, and Raphaëlle Sorba; Pr Dominique Turck (member of the French Society of Pediatrics and PI at the Lille University Hospital until 2023) and Dr Catherine Salinier (member of the French Association for Ambulatory Pediatrics) for their valuable engagement in the collaborative development, training and support for this interventional research; all dietitians for the implementation of the trial at the two study sites since 2017, namely: Yasmine Améziane, Solène Brun, Marjorie Daman, Marine Charlier, Emma Creps, Pauline Ditilyeu, Virginie Fourcaut, Noémie Jérôme, Sarah Mille, Amélie Siemiatkowski and Tiphaine Spas; all health care providers involved in the screening, monitoring or training at the two hospital maternity wards, especially Dr Sandy Hanssens, Karine Rogelet, Samantha Meyer, Hugues Bachelart, Nathan Storme, Florence Flamein, Aurore Dehon and Dr Pauline Devouge (PI at the Valenciennes Hospital); Céline Sardano-Garci, Faryal Harrar, and Uriel Makela at Inserm for their contribution to the data collection and/or the production of the participant newsletter; Coline Dumoulin, Françoise Fromageau, Thierry Couvert Leroy, and Marc Vannesson at the French Red Cross; and, last but not least, all participating families for their invaluable contribution to this research. The authors also thank Jo Ann Cahn for her help in preparing the manuscript.

## Authors’ contributions

SL leads the ECAIL trial (PI) and co-designed it with PH, LB, DD, DS, OS, BC, BLG, MAC, DL (PI at the Lille University Hospital); OS is responsible for managing the implementation of the ECAIL trial; PH monitored the data collection at the two study sites, provided part of the quantitative data analyzed in the present article and supervised the implementation team. DP, PS, and SL co-designed this interdisciplinary research; DP implemented the qualitative component, under the co-supervision of PS and SL; LV critically reviewed the qualitative findings; CLG contributed to data collection and conducted the quantitative analysis; DP and SL contributed to data collection and co-drafted the manuscript. All authors critically reviewed the manuscript; read and approved the final manuscript.

